# What contributes to pregnancy intendedness? Insights from the Dutch BluePrInt study using a conceptual hierarchical model

**DOI:** 10.1101/2025.06.18.25329857

**Authors:** Wieke Y. Beumer, Tessa J. Roseboom, Jenneke van Ditzhuijzen

## Abstract

**Background:** Pregnancy intentions are shaped by interrelated factors across individual, relational and societal contexts. This study employs a Conceptual Hierarchical Model (CHM) to examine pregnancy intentions among womxn and their partners.

**Methods:** Data were drawn from baseline measurements of the Dutch prospective BluePrInt study on unintended pregnancy. It included 911 participants (womxn and partners) who recently experienced an unexpected pregnancy, which they aborted or continued. Pregnancy intentions were assessed using the London Measure of Unplanned Pregnancy. Key variables included sociodemographics, social support, partner relationship, interpersonal violence, and mental health. A CHM guided multiple regression analyses, with additional analysis of sex differences.

**Results:** Findings indicated that educational attainment and social support were indirectly associated with pregnancy intendedness, while age, religiosity, cohabitation, and life satisfaction showed direct associations, with no evidence of a moderating effect of sex.

**Conclusions:** People who perceive their context as suitable for raising a child, who feel supported, and who have greater personal capacity to parent, perceive their initially unexpected pregnancy as more intended. Policies should promote social and relational stability universally, rather than targeting those experiencing unintended pregnancies. Reproductive counseling should address ambivalence and broader personal circumstances.

## Introduction

Unintended pregnancy is often considered a public health issue that should be prevented (1, 2). However, extensive evidence convincingly shows that social inequalities underlying unintended pregnancy (such as disparities in health access) are primary contributors to individual and public health problems (2–4). Although most previous studies focused on socioeconomic determinants of unintended pregnancy, such as age and socioeconomic position (SEP) (5–10), other factors also play a role, such as lack of social support, lower relationship quality, experiences of interpersonal violence and mental health issues (5, 6, 8–22). Notably, people may welcome their pregnancy, despite presence of any of these risks (23–25).

This highlights the need to move beyond individual determinants of unintended pregnancy, and consider how broader social and relational dynamics interact to shape pregnancy experiences. A conceptual hierarchical model (CHM) (26) adopts a socioecological approach (27) by studying how distal influences, such as SEP, affect unintended pregnancy through more immediate, proximal factors like mental health. By mapping these interrelated pathways, a CHM helps prevent underestimation of distal factors.

This CHM approach has been used to investigate determinants of unintended pregnancy in Malawi (6), but it has not yet been applied to other populations and contexts. While Hall and colleagues’ CHM (6) incorporated epidemiological determinants of unintended pregnancy, it did not account for psychosocial influences, which are equally important in understanding unintended pregnancy. Previous studies are limited in two additional ways. First, most have investigated pregnancy intentions using a binary distinction between intended and unintended pregnancies. These traditional measures have been critiqued extensively (2, 23, 25). Pregnancy intentions are captured better on a continuum, reflecting ambivalence, subconscious desires, or conflicting feelings. Research on unintended pregnancy as a multifaceted construct is limited (28), especially in countries with accessible abortion care. Second, previous studies primarily focused on womxn^1^, but it is also important to include partners’ perspectives in reproductive health research (29). Factors contributing to unintended pregnancy may differ for womxn and their partners^2^, though this remains understudied.

To address these research gaps, this study aims to hierarchically examine factors and their interrelations contributing to pregnancy intendedness, as a multifaceted, continuous construct, from the perspectives of womxn and their partners living in a context with relatively accessible abortion care (the Netherlands). Building on Hall and colleagues’ CHM analysis of unintended pregnancy (6), the current study’s CHM additionally incorporated psychosocial factors.

## Methods

### Study design

Data came from the first wave (December 2022 – August 2023) of the prospective BluePrInt study into unintended pregnancy in the Netherlands. It includes womxn and their partners who chose abortion (AB) or continuation of their pregnancy (CT).

Following consultations with professionals and experts, “unexpected pregnancy” was chosen as the recruitment term, as it was considered less stigmatizing and more in line with people’s experiences compared to alternatives like unintended.

AB participants were recruited from nine abortion clinics across diverse regions. In the Netherlands, more than 90% of abortions are performed in these clinics (up until 22 weeks gestation) (30). Recruitment of CT participants took place at 83 midwifery clinics nationwide (around eight weeks gestation).

People could sign up by scanning a QR code, either during their consult or in the waiting room. Additionally, CT participants were recruited through an online mailing around 14 weeks of gestation via their midwife, typically used for general pregnancy information. Partners were recruited via their pregnant partner.

The QR code linked to a webpage to sign up for the study. This included an information letter and video, and an online consent form. To avoid any potential decision-making interference, people could only participate after having an abortion or deciding to continue the pregnancy. People were eligible if they were older than 15 years. Further, CT participants were eligible if they scored at least two points on these criteria: (1) They were *not* trying to conceive before becoming pregnant (1 point); (2) they felt the pregnancy occurred at the *wrong* time (1 point) (3); their emotional response upon discovering the pregnancy was: *happy* (0 points), m*ixed feelings* (1 point), *unhappy* (1 point).

Once consent was provided and eligibility was confirmed, participants received a link to an online survey via email. The survey contained open- and closed ended questions. Most participants completed the questionnaire on their phones (20 minutes). Upon completion, they received a €10 gift card.

The study protocol was reviewed by the Medical Research Ethics Committee of the Amsterdam Medical Center under reference number W21_407, and received a waiver from the Medical Research Involved Human subjects Act (WMO).

### Participants

1,031 People filled out the online survey. For this specific analysis, participants with more than 50% missing values in the London Measure of Unplanned Pregnancy (LMUP) were excluded, following Barrett and colleagues’ advice (31). Hence, the final analyses contained 911 participants; 638 womxn (382 continued the pregnancy (CT), 256 had an abortion (AB)) and 273 partners CT-group: 142, AB-group: 131). Participants’ ages ranged from 16 to 49 years. CT participants completed the questionnaire around 14 weeks gestation. The majority of AB participants had a first trimester abortion (89.8%), and filled out the questionnaire afterwards. Power analyses indicated that this sample size was sufficient to answer our research questions (medium effect size, power >.9, *p* = 0.05).

### Measures

#### Outcome measure

An adapted version of the LMUP was used to measure pregnancy intentions (31). The LMUP includes six items on contraceptive use, preconceptional behavior (e.g., taking folic acid), partner communication, timing, desire for parenthood, and intention to become pregnant. Each item is scored 0 to 2, with a total score of 0 to 12; higher scores indicate a more intended pregnancy (Figure 1). The LMUP is valid and reliable (31, 32). The adapted LMUP that was used, has been found reliable in the Dutch context (current study’s Cronbach’s α = .762) (33).

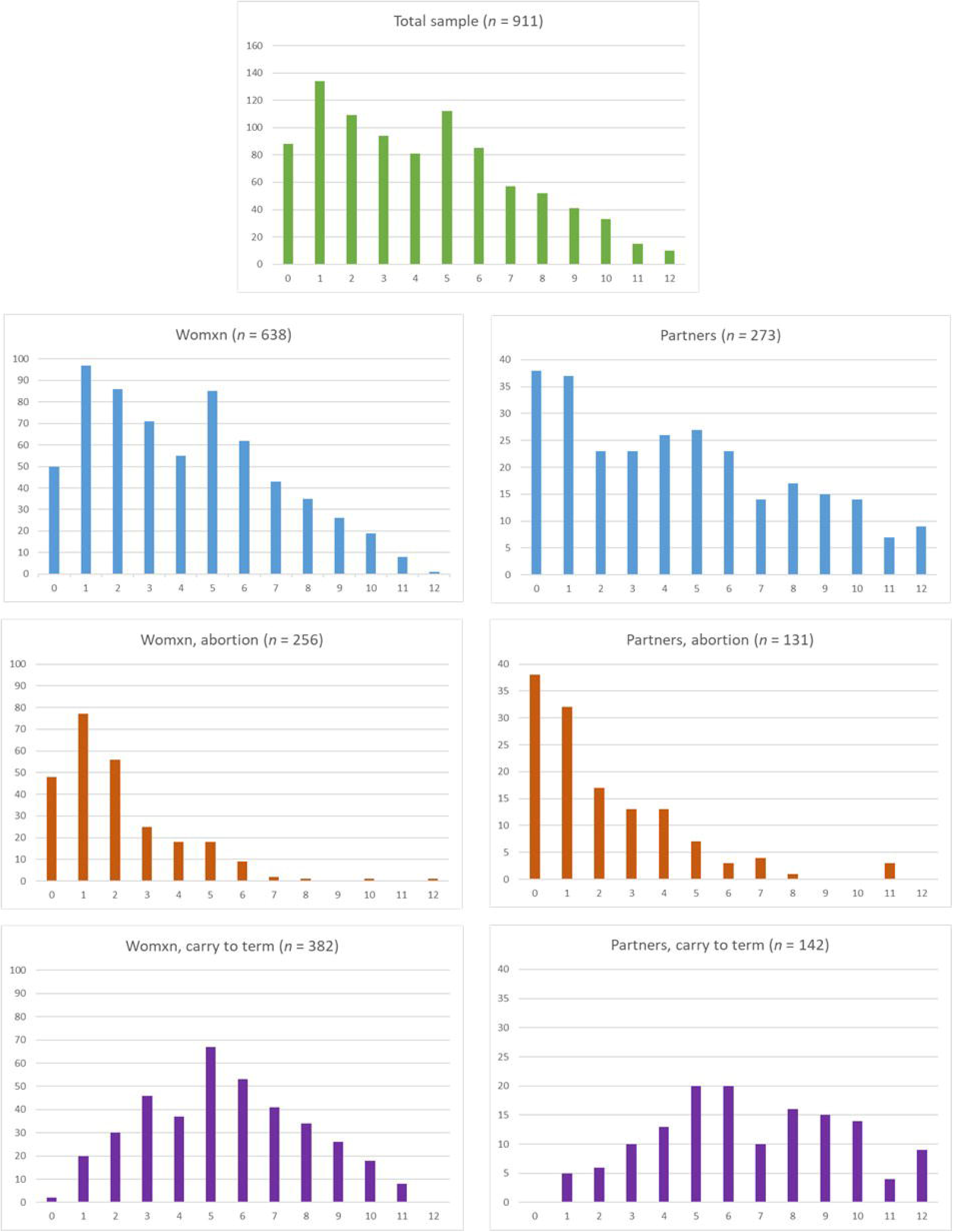

Participants could not leave survey questions blank but could answer with ‘Don’t know’ or ‘I’d rather not say,’ which were coded as missing. For participants with 1-3 missings, these were imputed with the mean (31).

### Independent variables

#### Socioeconomic

Education (theoretically vs. practically) and occupational status (having a job) were included as indicators of SEP.

#### Sociodemographic

These included: age, nationality (born in the Netherlands), religion (yes/no) and parity (nulliparous/multiparous).

#### Social support from family/friends

A subscale of the Multidimensional Scale of Perceived Social Support (MSPSS) measured social support from family/friends (34). The MSPSS consisted of five items, answered on a scale ranging from not at all (0) to completely (4). A mean score was calculated, with a higher mean indicating more perceived social support. The MSPSS is valid and reliable (34), also for the Dutch context (35), and in the current study (Cronbach’s α = .878).

### Partner relationship

#### Relationship status

We measured whether participants were in a romantic relationship, and were cohabiting with their partner. These were recoded into one variable (cohabiting vs not cohabiting).

#### Partner support

Another subscale of the MSPSS (34, 35) measured social support of a romantic partner, if present (no support (0) – a lot of support (4)). A mean score was calculated (Cronbach’s α = .932). People without a relationship were given a neutral score in the analyses.

#### Relationship satisfaction

Participants were asked ‘How happy are you in the relationship with your partner?’ (very unhappy (0) - very happy (4)). This measure was based on the shortened Relationship Assessment Scale (RAS-1) (36). People without a relationship were given a neutral score.

### Individual factors

#### Experiences with interpersonal violence

Participants indicated whether they experienced mental or physical childhood abuse (< 16 years old). These items were combined into any childhood abuse (yes/no). Further, participants indicated having experienced sexual victimization in their lifetime (yes/no).

#### Diagnosis of a psychiatric disorder

Participants reported whether they were ever diagnosed with a psychiatric disorder (based on the Lifetime Depression Assessment Self-report (LIDAS; (37)). We dichotomized answers into being diagnosed with one or more psychiatric disorders (yes/no).

#### Life satisfaction

Participants indicated how satisfied they were with their life on a 10-point scale, a higher score indicated more life satisfaction (38).

### Statistical analyses

All statistical analyses were performed in R studio (39). Collinearity was examined prior to selection of variables for inclusion in the hierarchical model selection process. Univariate associations between potential factors and the outcome were examined before developing a multiple regression model (*p* < .10). Since the total LMUP scores were not normally distributed (Figure 1), we computed the regression models with robust standard errors (40).

The CHM was used to guide multiple linear regression models. Factors were grouped into five levels (Figure 2). We started by analyzing the most distal level (socioeconomic), and worked down through increasingly more proximate levels. Factors higher in the hierarchy could influence unintended pregnancy either indirectly, through their effect on the factors in lower levels, or directly. At each level, variables were entered simultaneously. Variables with *p* > .10 were backward stepwise removed, starting with the highest *p*-value. Accepted variables were retained in the model, even if they later became non-significant. Lastly, we investigated sex differences, by adding sex as a moderator.

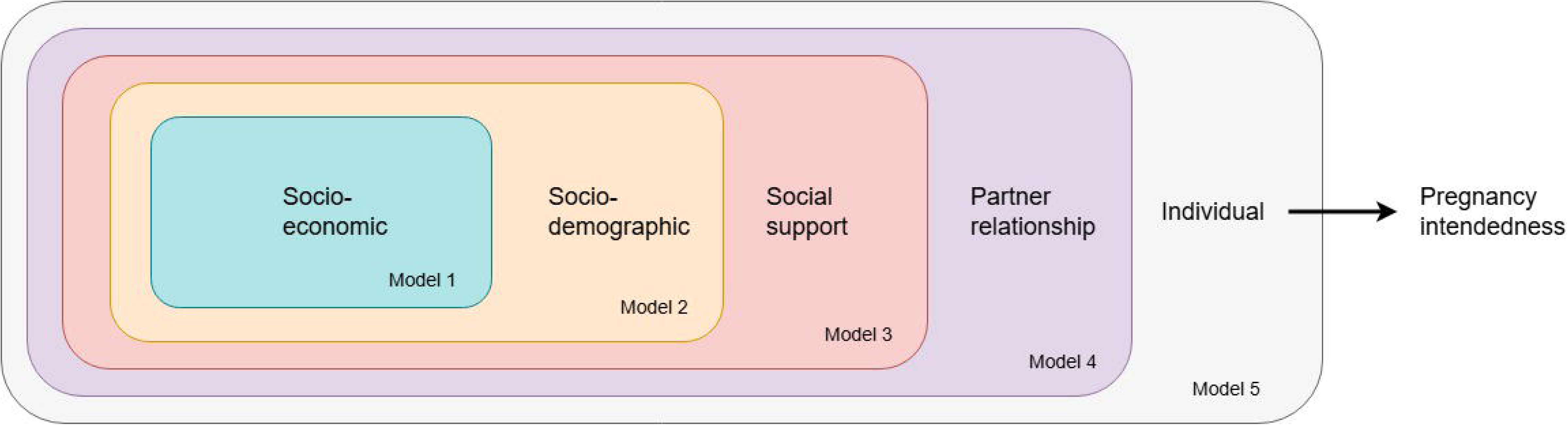

## Results

### Characteristics of participants

People in the AB-group generally reported lower pregnancy intendedness compared to people in the CT-group (Table I). Using suggested LMUP cut-points dividing pregnancies into ‘unplanned’, ‘ambivalent’ and ‘planned’ categories (31), most pregnancy intentions in the CT-group were ambivalent, while people in the AB-group mainly reported unintended pregnancies (Table I; Figure 1). The pregnancy intention scores of womxn and their partners showed similar distributions, also per pregnancy outcome (continuation or abortion).

**Table I.** Participant characteristics.

|  | **Womxn** |  |  | **Partners** |  |  |
| --- | --- | --- | --- | --- | --- | --- |
|  | **Total** | **Carry to term** | **Abortion** | **Total** | **Carry to term** | **Abortion** |
|  | *N* = 638 | *N* = 382 | *N* = 256 | *N* = 273 | *N* = 142 | *N* = 131 |
| **Unintended pregnancy** |  |  |  |  |  |  |
| LMUP (0-12) (*M*, *SD*) | 4.08 (2.8) | 5.44 (2.5) | 2.10 (1.9) | 4.48 (3.5) | 6.61 (2.8) | 2.13 (2.4) |
| LMUP categories (*N*, %) <sup>a</sup> |  |  |  |  |  |  |
| *Unintended* (score 0-3) | 304 (47.6) | 98 (25.7) | 206 (80.5) | 121 (44.3) | 21 (14.8) | 100 (76.3) |
| *Ambivalent* (score 4-9) | 306 (48.0) | 258 (67.5) | 48 (18.8) | 122 (44.7) | 94 (66.2) | 28 (21.4) |
| *Intended* (score 10-12) | 28 (4.4) | 26 (6.8) | 2 (0.8) | 30 (11.0) | 27 (19.0) | 3 (2.3) |
| **Socioeconomics** |  |  |  |  |  |  |
| Employed/studying | 560 (87.8) | 337 (88.3) | 223 (87.1) | 265 (97.1) | 138 (97.2) | 127 (96.9) |
| Education <sup>b</sup> |  |  |  |  |  |  |
| *Practical* | 394 (61.8) | 220 (57.6) | 174 (68.0) | 179 (65.6) | 82 (57.7) | 97 (74.1) |
| *Theoretical* | 242 (37.9) | 160 (41.9) | 82 (32.0) | 92 (33.7) | 59 (41.5) | 33 (25.2) |
| **Sociodemographics** |  |  |  |  |  |  |
| Born in the Netherlands (yes) | 596 (93.4) | 352 (92.1) | 244 (95.3) | 262 (96.0) | 134 (94.4) | 128 (97.7) |
| Age (*M*, *SD*) | 28.48 (6.1) | 29.46 (5.0) | 27.01 (7.1) | 28.56 (6.4) | 31.23 (5.3) | 2 (1.5) |
| 16-17 | 6 (0.9) | – | 6 (2.3) | 2 (0.7) | – | 70 (53.4) |
| 18-24 | 172 (27.0) | 56 (14.7) | 116 (45.3) | 83 (30.4) | 13 (9.2) | 54 (41.2) |
| 24-39 | 436 (68.3) | 316 (82.7) | 120 (46.9) | 173 (63.4) | 119 (83.8) | 5 (2.8) |
| >39 | 24 (3.8) | 10 (2.6) | 14 (5.5) | 15 (5.5) | 10 (7.0) |  |
| Religious | 149 (23.4) | 111 (29.1) | 38 (14.8) | 41 (15.0) | 33 (23.2) | 8 (6.1) |
| **Relational** |  |  |  |  |  |  |
| Relation (*N*, %) |  |  |  |  |  |  |
| *Single* | 55 (8.6) | 14 (3.7) | 41 (16.0) | 16 (5.9) | 3 (2.1) | 13 (9.9) |
| *In a relationship, not cohabiting* | 148 (23.2) | 51 (13.4) | 97 (37.9) | 67 (24.9) | 13 (9.2) | 54 (41.2) |
| *Cohabiting* | 421 (66.0) | 308 (80.6) | 113 (44.1) | 186 (69.1) | 123 (86.6) | 63 (48.1) |
| Parity (multiparous) | 227 (35.6) | 152 (39.8) | 75 (29.3) | 57 (20.9) | 40 (28.2) | 17 (13.0) |
| Social support of family/friends (0-4) | 3.15 (0.8) | 3.22 (0.7) | 3.06 (0.8) | 3.04 (0.7) | 3.01 (0.8) | 3.06 (0.7) |
| Social support of partner (0-4) | 3.34 (0.7) | 3.29 (0.7) | 3.4 (0.6) | 3.35 (0.5) | 3.32 (0.6) | 3.40 (0.5) |
| Happy in relationship (0-4) | 3.31 (0.7) | 3.29 (0.6) | 3.36 (0.7) | 3.44 (0.5) | 3.37 (0.5) | 3.52 (0.5) |
| **Individual** |  |  |  |  |  |  |
| Mental health diagnosis | 196 (30.7) | 130 (34.0) | 66 (25.8) | 39 (14.3) | 24 (16.9) | 15 (11.5) |
| Experience with sexual violence (ref = no) | 117 (18.3) | 70 (18.3) | 47 (18.4) | 3 (1.1) | 2 (1.4) | 1 (0.8) |
| Experience with physical/mental violence during youth (ref = no) | 94 (14.7) | 53 (13.9) | 41 (16.0) | 25 (9.2) | 11 (7.7) | 14 (10.7) |
| Satisfied with life (0-10) | 7.13 (1.6) | 7.55 (1.3) | 6.50 (1.8) | 7.68 (1.3) | 8.01 (1.9) | 7.32 (1.2) |

*Note* <sup>a</sup> LMUP categories were based on Barrett and colleagues cut-off points (31). <sup>b</sup> Educational attainment categories were based on the Dutch educational system. Practical education included: only primary school, VMBO, HAVO, VWO and MBO. Theoretical education included: HBO and WO.

### Hierarchical multivariable analyses

Multiple factors were not included in the final CHM (Table II). Some were not significantly associated with pregnancy intendedness in the univariate models (*p* > .10), such as employment status, interpersonal violence and parity (Table S1). Others were dropped as they were not significant in the CHMs (ethnicity, mental health diagnosis, happy with relationship).

**Table II.** Hierarchical multiple regression analyses with robust standard errors, modelling the factors contributing to greater pregnancy intendedness (measured with the LMUP).

|  | Model 1:<br>Socioeconomic |  |  |  | Model 2:<br>Sociodemographic |  |  |  | Model 3:<br>General social support |  |  |  | Model 4:<br>Partner relationship |  |  |  | Model 5:<br>Individual |  |  |  |
| --- | --- | --- | --- | --- | --- | --- | --- | --- | --- | --- | --- | --- | --- | --- | --- | --- | --- | --- | --- | --- |
| | $\beta$ | CI 95% | | *p* | $\beta$ | CI 95% | | *p* | $\beta$ | CI 95% | | *p* | $\beta$ | CI 95% | | *p* | $\beta$ | CI 95% | | *p* |
| Education (ref = theoretical) | -0.92 | 0.21 | -4.49 | <.001 | -0.16 | 0.21 | -0.75 | .453 | -0.09 | 0.22 | -0.4 | .687 | -0.04 | 0.21 | -0.19 | .852 | -0.04 | 0.21 | -0.2 | .845 |
| Religious (ref = no) |  |  |  |  | 1.06 | 0.23 | 4.61 | <.001 | 1.01 | 0.23 | 4.38 | <.001 | 1 | 0.23 | 4.41 | <.001 | 1.05 | 0.23 | 4.65 | <.001 |
| Age |  |  |  |  | 1.15 | 0.12 | 9.35 | <.001 | 1.11 | 0.12 | 9.03 | <.001 | 0.86 | 0.13 | 6.7 | <.001 | 0.81 | 0.13 | 6.41 | <.001 |
| Quadratic age |  |  |  |  | -0.02 | 0 | -8.76 | <.001 | -0.02 | 0 | -8.46 | <.001 | -0.01 | 0 | -6.43 | <.001 | -0.01 | 0 | -6.23 | <.001 |
| Social support family/friends (0-4) |  |  |  |  |  |  |  |  | 0.31 | 0.13 | 2.42 | .016 | 0.3 | 0.13 | 2.35 | .019 | 0.18 | 0.13 | 1.42 | .157 |
| Cohabiting (ref = no) |  |  |  |  |  |  |  |  |  |  |  |  | 1.51 | 0.22 | 6.98 | <.001 | 1.36 | 0.22 | 6.27 | <.001 |
| More satisfied with life (0-10) |  |  |  |  |  |  |  |  |  |  |  |  |  |  |  |  | 0.29 | 0.07 | 4.3 | <.001 |
| Adjusted R<sup>2</sup> goodness of fit |  |  |  | 0.021 |  |  |  | 0.143 |  |  |  | 0.147 |  |  |  | 0.193 |  |  |  | .209 |

The CHM results indicate that some variables directly affected pregnancy intendedness, and others indirectly through more proximal factors (Table II). Educational attainment was only significant in Model 1, suggesting an indirect effect via factors like social support. In contrast, religiosity and age (introduced in Model 2) consistently showed strong, direct associations with pregnancy intendedness. Religious individuals reported higher intendedness, and intendedness increased with age before declining around age 40. In Model 3, social support from family and friends emerged as a significant positive contributor to pregnancy intendedness. This association attenuated in Model 5, suggesting an indirect effect via individual factors like life satisfaction. Cohabitation, introduced in Model 4, was a strong and stable predictor, suggesting a direct effect.

Cohabiting individuals reported significantly higher pregnancy intendedness. Finally, life satisfaction, added in Model 5, was directly associated with higher pregnancy intendedness.

We also examined whether sex was a moderator in the final CHM. The inclusion of sex did not significantly improve model fit (Adjusted R² increased from .209 to .233). Additionally, there were no significant interaction terms, and sex did not significantly predict pregnancy intendedness (*p* = .308).

## Discussion

### Main findings

This study hierarchically examined interrelated factors that contribute to unintended pregnancy among both womxn and their partners, in a context where abortion care is relatively accessible (the Netherlands). We conceptualized unintended pregnancy as a continuous, multifaceted construct: hereafter called ‘pregnancy intendedness’. By using a Conceptual Hierarchical Model (CHM) from a socioecological perspective, we examined how distal factors affect pregnancy intendedness through more immediate, proximal factors, highlighting the importance of understanding unintended pregnancy within broader socioecological contexts. Unintended pregnancies do not occur in isolation, but are shaped by interactions between factors at individual, relational, and societal levels. Our results showed that people who live in a context they perceive suitable for raising a child, who feel supported, and have greater personal capacity to parent, perceive their initially unexpected pregnancy as more intended.

When zooming in on our results, the current study revealed several key factors contributing to people’s pregnancy intendedness. The most distal factor in our CHM, educational attainment, was indirectly associated with pregnancy intendedness through more proximate factors. People with lower educational attainment generally face more socioeconomic challenges (41), which may influence pregnancy intendedness indirectly via relational and individual factors. Moving on to more proximate factors, pregnancy intendedness tended to increase with age, up until in late thirties and decreased after 40. This trend implies that people within the typical ‘reproductive age’ range perceive their personal life stage as more suitable for raising a child (42). Another important factor was religiosity, as religious people reported higher pregnancy intendedness, potentially reflecting cultural values or beliefs in divine planning (24). Furthermore, cohabiting individuals and those who perceived greater social support reported greater pregnancy intendedness, as did those with higher life satisfaction. These factors may offer a sense of stability, making an unexpected pregnancy feel more manageable. However, due to the study’s cross-sectional design, the direction of these relationship remains unclear.

Contrary to expectations, several factors were not associated with pregnancy intendedness. Mental health diagnoses showed no significant association, even in sensitivity analyses using alternative self-reported measures, possibly due to self-report bias. Parity was also not linked to pregnancy intendedness. This may be due to the study’s comparison of nulliparous and parous individuals without accounting for the number of children. Additionally, experiences with interpersonal violence were not associated with pregnancy intendedness, potentially related to differences in measurement from prior studies (6, 9, 11, 18, 43).

### What is already known on this topic

The findings of the current study align with Macleod’s pregnancy supportability framework (25), which posits that someone perceives their pregnancy as more supported based on a combination of their capacities to carry a pregnancy, enabled or constrained through individual circumstances and broader social support systems. This framework highlights the importance of examining how interrelated factors across societal, relational and individual contexts contribute to people’s pregnancy intentions.

Most prior studies focused on sociodemographic determinants of unintended pregnancy, for instance showing that higher educational attainment and older age are associated with higher pregnancy intendedness (5–8, 43, 44). Previous studies also showed that those who cohabit, report strong relationships, or perceive greater social support report higher intendedness (5–7, 9, 10, 13–16, 40, 43, 44). Conversely, people with children, mental health issues, or experiences of interpersonal violence often report lower intendedness (6, 8, 9, 11, 12, 16, 18, 20, 21, 43). However, findings on mental health are mixed, with some studies, like Tenkky et al. (45), finding no association.

### What this study adds

This study is the first to examine the underlying factors of unintended pregnancy among both womxn and their partners in greater depth. It revealed similar associations for womxn and their partners. Including partners offered valuable additional insights, revealing shared dynamics and perspectives that would have remained hidden if only womxn had been studied. A further contribution of this study to the existing literature is its multifaceted conceptualization of pregnancy intentions, combined with the application of a CHM grounded in a socioecological framework, which enabled a nuanced analysis of how distal societal factors influence pregnancy intendedness through relational and individual pathways. Furthermore, this study was among the first to investigate pregnancy intentions of both people who chose abortion or continuation of their unexpected pregnancy. Our data showed that in a context with relatively accessible abortion care, most people who opted for an abortion experienced a highly unintended pregnancy, while those who continued their pregnancy often reported ambivalent intentions. However, there was considerable diversity in pregnancy intentions regardless of the decision made. Beyond intentions, other aspects (such as emotions and a ‘sense of knowing’) play a crucial role in the decision to have an abortion or not (46, 47). The factors related to pregnancy decision-making in the BluePrInt study are reported elsewhere (48).

These findings have important implications for public health. Similar to other public health interventions (49), efforts to reduce unintended pregnancy often narrowly focus on individual behavior, while overlooking the need for structural policies that address underlying social inequities. These interventions typically aim to improve individuals’ ability to plan pregnancies, by recommending effective contraceptive use. While access to contraception is crucial for reproductive justice, interventions that target individual contraceptive behavior mainly emphasize personal responsibility (25). Such approaches can be stigmatizing, overlook the structural and cultural factors that shape reproductive experiences, and are imbued with normative beliefs that unintended pregnancies are negative events that should be prevented, despite (scientific) critiques of these beliefs (2, 25). They also fail to accommodate ambivalent, indifferent, or fluctuating pregnancy desires (23), and ignore that planning a pregnancy is irrelevant or unattainable for some people due to cultural beliefs or socioeconomic constraints (24, 50).

In line with earlier recommendations (2, 23, 25, 51), we propose that policies and healthcare providers develop people-centered approaches connecting more closely with the complexity and diversity in people’s reproductive experiences, emotions and desires. This includes ensuring access to abortion care and contraceptive counseling with both womxn and their partners, while addressing broader social inequalities, such as improving gender equality (52), that shape reproductive outcomes.

### Limitations

Owing to the observational design, no causal relationships could be established. Further, assessing pregnancy intention antepartum or shortly after abortion remains retrospective, not capturing pre-conception intent. People continuing their pregnancy often report higher intention over time (53, 54), which might have influenced underreporting of unintended pregnancy in the current study. Moreover, our sample underrepresented people born outside the Netherlands, those with lower educational attainment, and with a religious background (55–57). This underrepresentation may have skewed our results, potentially underestimating the impact of these factors. Lastly, although the CHM was literature-based, the order of levels was subjective. However, sensitivity analyses confirmed it did not affect the results.

## Supporting information

Supplementary Tabel 1

## Data Availability

All data produced in the present study are available upon reasonable request to the corresponding author.

## Acknowledgements

We would like to extend our gratitude to all the participants who took the time to respond to our BluePrInt survey. We are also appreciative of everyone who assisted in recruiting participants.

Our sincere thanks go to the experience experts and professional advisors whose thoughtful and meaningful insights greatly enriched this study. In particular, we would like to acknowledge Annemarie Reilingh for her valuable contributions to our discussions on unintended pregnancy within the BluePrInt study. Finally, we wish to express our profound gratitude to Prof. Dr. Ellen Laan, who played a pivotal role in the development of the grant proposal for this study. Her involvement and support were deeply appreciated. We are saddened by her passing and honor her lasting impact on this work.

## Footnotes

1 This also includes transgender men and nonbinary individuals with the capacity for pregnancy.

2 We define ‘partners’ as the people who are involved in the pregnancy either as a romantic partner (regardless of their gender and/or sex), or a non-romantic, but sexually/biologically involved partner.

