## Supplementary Tabel 1 for "What contributes to pregnancy intendedness? Insights from the Dutch BluePrInt study using a conceptual hierarchical model"

**Table S1.** Univariate linear regression with robust standard errors analyzing factors contributing to higher pregnancy intendedness (measured with the LMUP).

|  | β | 95% BI | | *p* |
| --- | --- | --- | --- | --- |
| **Socioeconomics** |  |  |  |  |
| Education (ref = theoretical) | -0.92 | -1.33 | -0.52 | <.001 |
| Having a job (ref = no) | 0.47 | -0.19 | 1.13 | .210 |
| **Sociodemographics** |  |  |  |  |
| Country of birth (ref = born in the Netherlands) | 1.05 | 0.18 | 1.91 | .018 |
| Age | 0.1 | 0.07 | 0.13 | <.001 |
| Religious (ref = no) | 1.24 | 0.78 | 1.7 | <.001 |
| **Relational** |  |  |  |  |
| Cohabiting (ref = no) | 2.13 | 1.77 | 2.5 | <.001 |
| Parity (ref = nulliparous) | 0.13 | -0.28 | 0.55 | .545 |
| Social support of family/friends (0-4) | 0.52 | 0.29 | 0.75 | <.001 |
| Social support of partner (0-4) | 0.4 | 0.25 | 0.55 | <.001 |
| Happy in relationship (0-4) | 0.32 | 0.06 | 0.58 | .021 |
| **Individual** |  |  |  |  |
| Mental health diagnosis | 0.58 | 0.15 | 1.01 | .011 |
| Experience with sexual violence (ref = no) | 0.3 | -0.28 | 0.87 | .318 |
| Experience with physical/mental violence during youth (ref = no) | 0.01 | -0.59 | 0.6 | .984 |
| Satisfied with life (0-10) | 0.49 | 0.37 | 0.61 | <.001 |
